# Environment, animal hosts and human activity on predicting space-time variations of Lyme Borreliosis incidence in France: a Bayesian two-part model

**DOI:** 10.1101/2022.05.30.22275741

**Authors:** Wen Fu, Camille Bonnet, Alexandra Septfons, Julie Figoni, Jonas Durand, Pascale Frey-Klett, Denis Rustand, Benoît Jaulhac, Raphaëlle Métras

**Affiliations:** Sorbonne Université, INSERM, Institut Pierre Louis d’Épidémiologie et de Santé Publique, Paris, France; Santé publique France, Saint-Maurice, France; Tous Chercheurs Laboratory, Université de Lorraine, INRAE, IAM, Champenoux, France; Statistics Program, Computer, Electrical and Mathematical Sciences and Engineering Division, King Abdullah University of Science and Technology (KAUST), Thuwal, Kingdom of Saudi Arabia; French National Reference Center for Borrelia, Hôpitaux Universitaires de Strasbourg, Strasbourg, France; Institut de Bactériologie, Fédération de Médecine Translationnelle de Strasbourg, University of Strasbourg, UR7290, ITI InnoVec, 3 rue Koeberlé, Strasbourg, France; Centre for Mathematical Modelling of Infectious Diseases, London School of Hygiene & Tropical Medicine, Keppel Street, London, UK; Department of Infectious Disease Epidemiology, London School of Hygiene & Tropical Medicine, Keppel Street, London, UK

## Abstract

**Background:** Lyme Borreliosis (LB) is the most widespread hard tick-borne zoonosis in the Northern Hemisphere and shows a seasonal pattern. Existing studies in Europe mainly focused on acarological risk assessment, with very limited investigations exploring human LB occurrence. We aimed to highlight areas and seasons of higher risk for LB occurrence in mainland France, integrating information on meteorological, environmental, animal hosts and human exposure to quantify the associated spatial and temporal risk factors.

**Methods:** We fitted 2016–19 French LB surveillance data to a two-part spatiotemporal statistical model, defined with binomial and gamma distributions, to explore the factors associated with the presence and increased LB incidence. Shared spatial and temporal random effects were specified using a Besag-York-Mollie model and a seasonal model, respectively. Coefficients were estimated in a Bayesian framework using integrated nested Laplace approximation. Projections and data for 2020 were used for model validation.

**Findings:** LB presence was associated with a high vegetation index (≥0·6). LB incidence increased in areas highly suitable for deer (≥80% cover per area), with mild soil temperatures (10–15°C) in the season preceding the onset, moderate air saturation deficits (3–5 mmHg), and higher proportion of tick bite reports. Prediction maps showed a higher risk of LB in spring and summer (April-September). Substantial geographical variation in LB incidence was found. Higher incidence was reported in parts of eastern, midwestern, and southwestern France.

**Interpretation:** This is the first national-level assessment of seasonal human LB occurrence in Europe allowing to disentangle factors associated with LB presence and increased incidence. This model illustrates a spatial integrated analysis of meteorological, hosts, and anthropogenic factors for a zoonotic and vector-borne infection of major public health concern, and can be used as a reference model to be calibrated in other LB-affected areas.

**Funding:** WF is funded by a Sorbonne University PhD fellowship, JD is supported by a grant overseen by the French National Research Agency (ANR) as part of the «Investissements d’Avenir» program (ANR-11-LABX-0002-01, Lab of Excellence ARBRE).

## Introduction

Vector-borne diseases are prioritized by World Health Organization (WHO) as a growing global public health concern, with the expected goal of reducing the incidence of cases worldwide by 60% in 2030 compared to 2016.^1^ Ticks are able to transmit a large variety of zoonotic infectious agents amongst animals, and from animals to humans, causing multiple infections in endemic areas, and raising concern in currently disease-free areas.^2^ In the Northern Hemisphere, Lyme borreliosis (LB), caused by the spirochete *Borrelia burgdorferi* sensu lato (*B. burgdorferi* s.l.) species complex and transmitted by hard ticks *Ixodes*, is the most prevalent tick-borne zoonosis.^3^ LB infection may be asymptomatic or manifests as an erythema migrans (EM, typical Lyme skin rash).^4^ In rare cases, the bacteria may spread to other organs, such as joints, nervous system, and heart.^4^ In Western Europe, the estimated annual incidence of LB is approximately 22/100,000 inhabitants, with wide variation across geographical regions, ranging from 464/100,000 in southern Sweden to 0·001/100,000 in Italy.^5^

In endemic regions, the persistence of *B. burgdorferi* s.l. is permitted by a complex epidemiological cycle at the interface between *Ixodes* ticks and animals, which act as reproductive or amplifying hosts.^6^ A large number of animals can provide a blood meal for *Ixodes* ticks (including birds, reptiles, and mammals), only few host species in Europe can act as a reservoir for *B. burgdorferi* s.l. (such as Eurasian blackbird, yellow-necked mouse, and bank vole).^7^ The presence and abundance of infected ticks therefore depend on the distribution of animal hosts, as well as suitable climates and vegetation habitats, allowing host-seeking, molting, and the completion of *Ixodes’* life cycle (egg, larvae, nymph and adult).^8^ Spillover to humans can occur through the bite of infected ticks, during outdoors activities. Understanding the spatial heterogeneity of LB incidence in humans therefore necessitate to account for environmental, meteorological factors and animal hosts’ distribution that can influence the presence and abundance of infected ticks, as well as humans’ exposure to infectious tick bites.

Numerous studies conducted in Europe have focused on acarological risk assessment and have been realized in fragmented, tick-friendly environments.^9^ A large number of biotic and abiotic factors related to the ecology of *Ixodes (I*.*) ricinus* (the primary vector for human LB in Europe) have been tested individually or synergistically, but their limited spatial scales, their varying protocol designs and different analytical applications make results difficult to compare and generalize.^9^ Among these, climatic factors such as temperature, humidity and saturation deficit have found to be good proxies for the host-seeking activity of nymphs.^10,11^ Biotic factors such as vegetation indices and rodent population have also been shown to be associated with tick abundance and infection rates.^12^ Anthropogenic data characterizing human outdoor activity or human exposure to infected tick is challenging to collect.^8^ Yet, recent citizen science research have shown promising results on characterizing human exposure to tick bite.^13^ In addition, large-scale exploration of these above variables in relation to human LB occurrence in a comprehensive framework has not yet been done. Addressing this research gap is necessary to better understand the impact of climate, environment, and animal hosts on human health and to assist policy makers in developing measures to prevent and control LB or other *Ixodes*-borne diseases.

In France, LB is a growing public health concern and has raised the attention of the French national government.^14^ Since 2009, the national sentinel network (Réseau Sentinelles, RS) monitors LB incidence.^15^ To date, LB cases have been reported in all regions of France (French administrative divisions, level 1), with a clear seasonal pattern, peaking between May and August.^16^ Reported incidence rates vary largely between the areas, ranging from 251 cases (95% CI [127; 375]) per 100,000 inhabitants in Alsace to 12 cases (95% CI [0; 35]) per 100,000 inhabitants in Basse-Normandie in 2019.^15^

This study maps areas and seasons at higher risk of LB in mainland France at a fine spatial resolution and quantifies the associated spatial and temporal risk factors. For this, we fitted 2016–19 national surveillance data to a two-part statistical model in a Bayesian framework, to estimate the association between LB incidence and a range of spatio-temporal covariates assumed to be potential risk factors (including meteorological, environmental, animal host, and anthropogenic factors). We used 2020 data for model validation.

## Materials and methods

### Study design and projection

We divided mainland France into a grid of 1,753 cells, each with a size of approximately 0·2 × 0·2 decimal degrees (dd) (≈22km^2^), as the unit of spatial analysis. Time was expressed in discrete units of three months (or quarters), divided in winter (January to March), spring (April to June), summer (July to September), and autumn (October to December).

All spatial data were rasterized and resampled to match the resolution of the grid cells and projected to the World Geodetic System (WGS84) system.

### LB surveillance data

We produced quarterly incidence rate by department (administrative division level 2) by aggregating departmental LB weekly incidence (/100,000 inhabitants) from national surveillance data, for the period 2016–19 (https://www.sentiweb.fr). These incidence estimates were generated from LB cases reported by general practitioners participating in continuous surveillance in primary care.^17^ The case definition used was based on the guidelines of the European Study Group on Lyme Borreliosis (ESGBOR), with LB cases diagnosed by the presence of an erythema migrans (EM), or of at least one disseminated manifestation, confirmed by ELISA and Western blot tests.^18^

National maps at our study resolution (0·2 × 0·2 dd) were generated from departmental incidence by spatial interpolation using ordinary kriging. The centroid of each department was used as the geographical coordinate to define the semivariogram function, that is, the semi-variance of incidence rates from two different locations in relation to the distance between them.^19^ A spherical model was used to fit the experimental semivariogram and incidence values were estimated for each grid cell. We mapped the kriged LB incidence for each quarter and used this as outcome data in the model.

### Space-time model specification

Let *Y*_*ij*_ be the kriged LB incidence rate at *x*_*i*_^*th*^ grid cell and *t*_*j*_^*th*^ quarter, including zero and positive continuous values. We used a two-part model that decomposed the distribution of *Y*_*ij*_ into a binary outcome (absence *versus* presence) fitted with a logistic mixed effects model and a continuous outcome (all positive values) fitted with a gamma mixed effects model. Spatial random effects and seasonal variation are shared between the models. For both parts of the model, covariates (time-varying or fixed) were preselected as potential risk factors according to LB eco-epidemiology, and examined independently. Selection of covariates are detailed in Table 1 and presented hereinafter. The complete model is defined as follows:

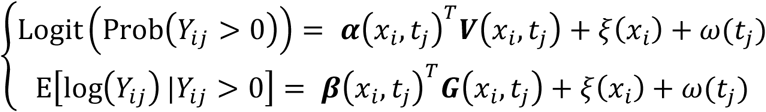

**Table 1.** Covariates selected for the two-part model, along with their descriptions and hypotheses with regards to LB occurrence.

|  |  |  |
| --- | --- | --- |
| **Variable** | **Description** | **Hypotheses** |
| **Logistic model (LB presence versus absence)** |  |  |
| Environmental and *Borrelia* reservoir factors associated with infected ticks, correlating with the spatiotemporal presence of LB in humans. |  |  |
| Normalized difference vegetation index (NDVI) | NDVI values reflect the photosynthetic activity of green plants and are aggregated by season. | NDVI is recommended as an important proxy for predicting tick distribution, reflecting the spatial and temporal dynamics of tick activity.<sup>23</sup> Higher NDVI values are positively correlated with the number of questing nymphs.<sup>23</sup> |
| Indices of rodent species richness | Predicted average number of species in the field (five species: *Apodemus agrarius*/ *flavicollis*/ *sylvatica*, *Microtus arvensis*, *Myodes glareolus*) | The number of potential host species may influence the spread of the pathogen, by increasing or decreasing contact with other hosts and thus affecting the distribution of the disease or its vectors.<sup>24</sup> |
| **Gamma model (Increase of LB incidence conditional on LB presence)** |  |  |
| Animal host, meteorological and anthropogenic factors influence tick density, questing activity, and the frequency of human-tick contact, driving LB risk increase or decrease. |  |  |
| Index of deer presence | Percentage (%) of suitable habitat for roe deer *Capreolus capreolus* or red deer *Cervus elaphus* presence | The presence of high deer density is associated with increased tick abundance, suggesting elevated risk of LB.<sup>9</sup> |
| Soil temperature (ST) | Average maximum soil temperature per quarter (°C, level 1:0–7cm) | As spring warms, ticks start questing activity, ie, contact with hosts and blood-feeding, and temperature thresholds (average maximum temperature) have been observed at 10°C for larvae and 7–8°C for nymphs, with peaks occurring at 15–17°C.<sup>11,25</sup> |
| Saturation deficit (SD) | A mixture index calculated from average maximum air temperature and humidity, averaged by quarter | SD could explain important changes in the number of questing ticks, mostly occurring between 2 and 7 mmHg and initiating a decline when SD>5 mmHg.<sup>26</sup> Higher SD values are associated with a decrease in the number of questing ticks.<sup>27</sup> |
| Rainless days | Cumulative number of rainless days per quarter. A rainless day is defined as a daytime period (6am–6pm) when the average depth of precipitation covering the entire surface is less than 1mm per hour | Precipitation is assumed to be negatively correlated with human physical activity.<sup>28</sup> We used this to explore the likelihood of people engaging in outdoor activities and the incidence of LB. |
| Frequency of tick bite reports | Proportion of quarterly tick bite reports per department, adjusted for their population weights | People may develop LB infection after being bitten by ticks.<sup>29</sup> |

Where ***V*** and ***G*** are vectors of covariates at location *x*_*i*_ at quarter *t*_*j*_ associated with the outcome in binary part and continuous part, respectively. ***α*** and ***β*** are vectors of coefficients for the covariates and we assigned a Gaussian prior with mean zero and precision 0·001. Spatial random effect *ξ*(*x*_*i*_) was specified using a Besag-York-Mollie model. It consists of an intrinsic conditional autoregressive model for spatially structured effects *u*_*i*_, and an independent identically distributed (iid) Gaussian model for spatially unstructured effects *ν*_*i*_.^20^

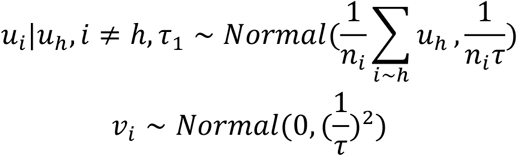

where *i* ∼ *h* indicate that two locations *i* and *h* are first-order neighbours, *n*_*i*_ is the number of neighbours of location *i*, with *τ* being the precision parameter.

Seasonal variation *ω*(*t*_*j*_) can be represented as a set of random vectors ***ω***=(*ω*_1_, *ω*_2_,…, *ω*_*j*_) with periodicity *s*. The density for ***ω*** is derived from the *j* − *s* + 1 increments as

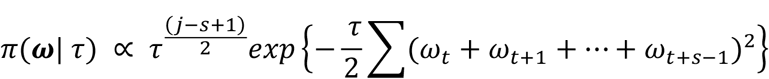

Here s=4 denotes the number of quarters in a year and j=16 denotes the total number of quarters in 2016–19. We set the priors for precision parameter *τ* as Gamma (1, 0·00005).

### Inference framework and parameter estimation

The fixed and random effects coefficients were estimated from data by Bayesian inference. We used the integrated nested Laplace approximation (INLA) method to approximate the posterior marginal distribution of the parameters of the two-part model. Compared with sampling-based methods such as MCMC, INLA can achieve Bayesian inference more efficiently and accurately.^20^ All analyses were performed in R version 4.0.5 using the INLA package.^21^

### Model selection and validation

Univariable and multivariable analyses were conducted; covariates were categorised by biological relevance (Table 1) and quartiles. We used the widely applicable information criterion (WAIC) to select the preferred multivariable model.^22^ Cross-validation was performed using data from 2020. Projection maps and the empirical distribution of the probability integral transform (PIT) were produced to assess the predictive performance of our model.

### Environmental and animal data

We used Copernicus Global Land Service version 2 (300m resolution) and version 3 (1km resolution) products, for the period 2016 to December 2020. All files were downloaded in NETCDF format, and summarized by quarter.

For the animal hosts data, we used two deer presence layers (Roe deer *Capreolus* or Red deer *Cervus elaphus*) and merged into one single layer, and another layer of five rodent species. These files are predicted values derived from three different spatial models conducted by the Environmental Research Group (ERGO). Detailed descriptions can be found in reference to their literature. The animal data used here are fixed in time.

### Meteorological data

Soil temperature (ST, level 1: 0-7 cm), air temperature (AT) and relative humidity (RH) above ground level, and precipitation (PP) were retrieved from the Copernicus climate data store using hourly values from the ERA5 and ERA5-Land Reanalysis datasets for the years 2016 to 2020. The grid data were downloaded in NETCDF format. For each grid cell, we aggregated the maximum mean values of ST, AT, and RH by quarter. Saturation deficits (SD) was calculated using the formula: SD=(1-RH/100)*4·9463*exp (0·0621*AT).^27^ PP was used to calculate the number of rainless days per quarter. All processing of meteorological data was performed using raster, ncdf4, ecmwfr and keyring packages in R version 4.0.5.

### Anthropogenic data on tick bites

Signalement TIQUE is a smartphone application and website developed by the national CiTIQUE participatory research programme to better understand the frequency and extent of tick-hosts contact, tick ecology, and infection status across mainland France (www.citique.fr). The application was launched in July 2017 and updated in May 2020.

A total of 33,531 human tick bite reports were included in the analysis. For each report, we extracted the information on reporting dates and GPS position (WGS84). Since information before July 2017 was not available, we made a compromise by retaining the average seasonal variation for each department throughout the study period in the model (appendix pp 2).

## Results

In Kriging interpolation, the semi-variogram fitted by the spherical model shows that the incidence distribution displayed spatial autocorrelation up to 110 km (Figure S1, appendix pp3). Corresponding smoothed quarterly maps of LB incidence for the 2016–19 years, used as our outcome variable are presented in appendix pp 3-5, exhibiting an increased incidence in spring and summer, mainly in the northeastern and eastern areas of the country (Figure S2, p4). Description of covariates included in this study is shown in appendix (pp 6-8).

The results of the preferred two-part model with the smallest WAIC value are reported in Table 2, and detailed as follows. The results from the first part of the model (logistic model), show that areas with higher vegetation activity, ie, NDVI values above and equal to 0·6, had a 42% higher odds of LB presence than areas with lower NDVI (OR=1·42, 95% CrI [1·31–1·53]). Conversely, the rodent species richness index was negatively correlated with LB presence, with the odds of LB occurring being 27% lower in areas with increased rodent species (OR=0·73, 95% CrI [0·67–0·79]). In the second part of the model (gamma model), areas with more than 80% cover of suitable habitat for deer had 1·12 times higher risk of LB compared to areas with less than 40% cover (RR=1·12, 95% Crl [1·03–1·22]. The risk of LB in the quarter Q increased by 1.18 in areas with mild soil temperature (ST, ranging between 10°C and 15°C) in the previous quarter (Q-1), but decreased for temperatures higher than 22°C (ST 10–15°C, RR=1·18, 95% Crl [1·10–1·25]; ST> 22°C, RR=0·77, 95% Crl [ 0·70–0·84]). Similarly, the risk of LB increased by 1·17 in areas with air saturation deficit (SD) between 3 and 5 mmHg compared with areas with SD below 3 mmHg (RR=1·17, 95% Crl [ 1·10–1·24]). This effect was reversed with increasing SD values; with the risk of LB halved in areas with SD higher than 9 mmHg (RR=0·56, 95%Crl [0·51–0·62]). Finally, the frequency of tick bite reports was also an important predictor of LB risk, with an increased risk of LB in locations and seasons where a higher proportion of bites were reported (Table 2).

**Table 2.**
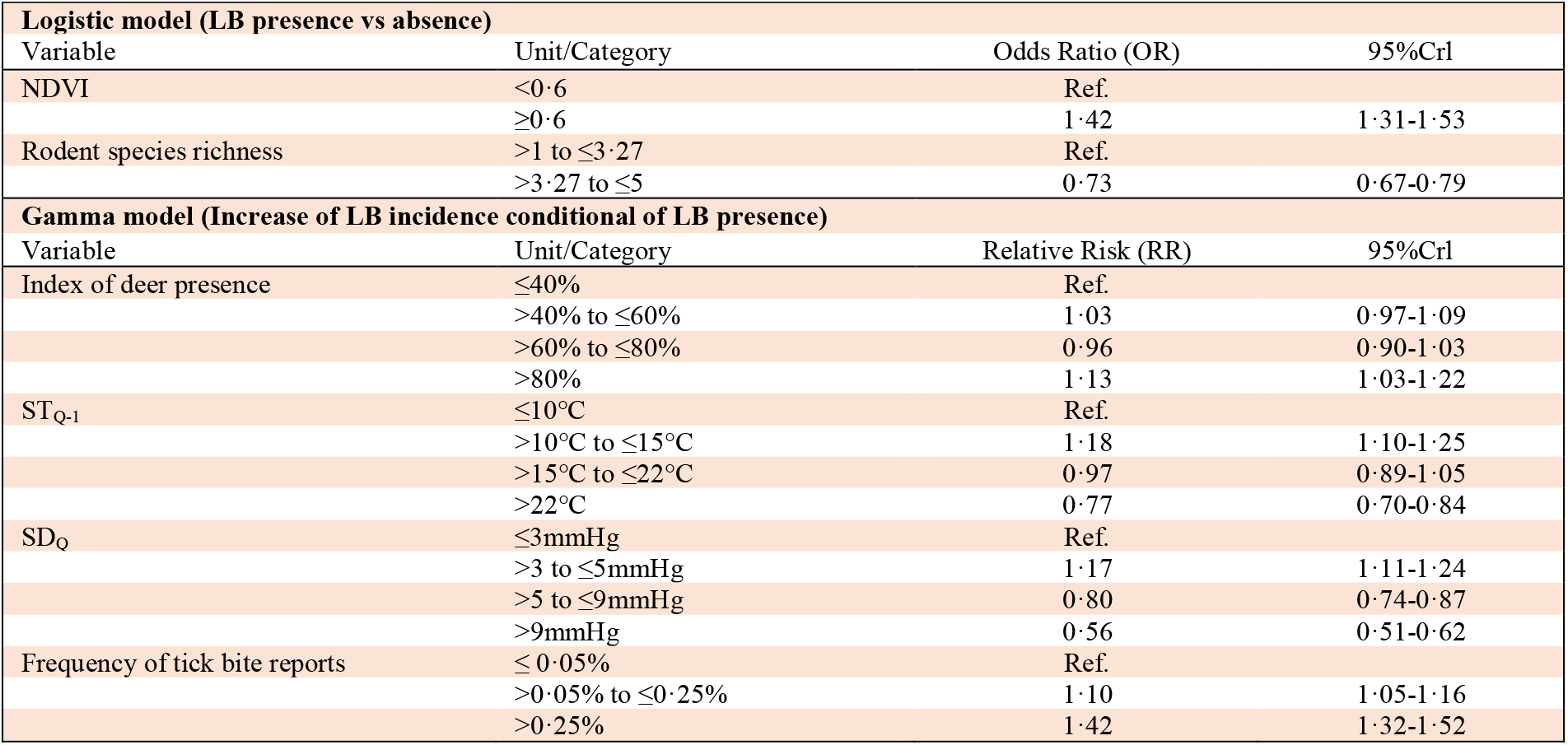
Results of two-part multivariable model for the years 2016-2019. Exponentiated the mean value of the posterior marginal distribution of coefficients and their 95% credibility intervals (Crl).

| **Table 2 Results of two-part multivariable model for the years 2016-2019. Exponentiated the mean value of the posterior marginal distribution of coefficients and their 95% credibility intervals (CrI).** |  |  |  |
| --- | --- | --- | --- |
| **Logistic model (LB presence vs absence)** |  |  |  |
| Variable | Unit/Category | Odds Ratio (OR) | 95%CrI |
| NDVI | <0.6 | Ref. |  |
|  | ≥0.6 | 1.42 | 1.31-1.53 |
| Rodent species richness | >1 to ≤3.27 | Ref. |  |
|  | >3.27 to ≤5 | 0.73 | 0.67-0.79 |
| **Gamma model (Increase of LB incidence conditional of LB presence)** |  |  |  |
| Variable | Unit/Category | Relative Risk (RR) | 95%CrI |
| Index of deer presence | ≤40% | Ref. |  |
|  | >40% to ≤60% | 1.03 | 0.97-1.09 |
|  | >60% to ≤80% | 0.96 | 0.90-1.03 |
|  | >80% | 1.13 | 1.03-1.22 |
| ST<sub>Q-1</sub> | ≤10°C | Ref. |  |
|  | >10°C to ≤15°C | 1.18 | 1.10-1.25 |
|  | >15°C to ≤22°C | 0.97 | 0.89-1.05 |
|  | >22°C | 0.77 | 0.70-0.84 |
| SD<sub>Q</sub> | ≤3mmHg | Ref. |  |
|  | >3 to ≤5mmHg | 1.17 | 1.11-1.24 |
|  | >5 to ≤9mmHg | 0.80 | 0.74-0.87 |
|  | >9mmHg | 0.56 | 0.51-0.62 |
| Frequency of tick bite reports | ≤0.05% | Ref. |  |
|  | >0.05% to ≤0.25% | 1.10 | 1.05-1.16 |
|  | >0.25% | 1.42 | 1.32-1.52 |

To visualise our results, we generated projection maps for each part of the models. Figure 1A-1P displays the predicted probability of LB presence for each quarter during 2016–19. We observed a seasonal pattern of disease occurrence, with higher probabilities in spring and summer, in almost every region of the country. Conversely, the probability of occurrence distribution in autumn and winter was much lower but presented important geographical heterogeneity, with high-risk areas concentrated in the Grand Est (GE), Bourgogne-Franche-Comté (BFC), and Auvergne-Rhône-Alpes (ARA) regions in eastern France; and in the Nouvelle Aquitaine (NA), and Occitanie (OT) regions in midwestern and southwestern France.

**Figure 1.**
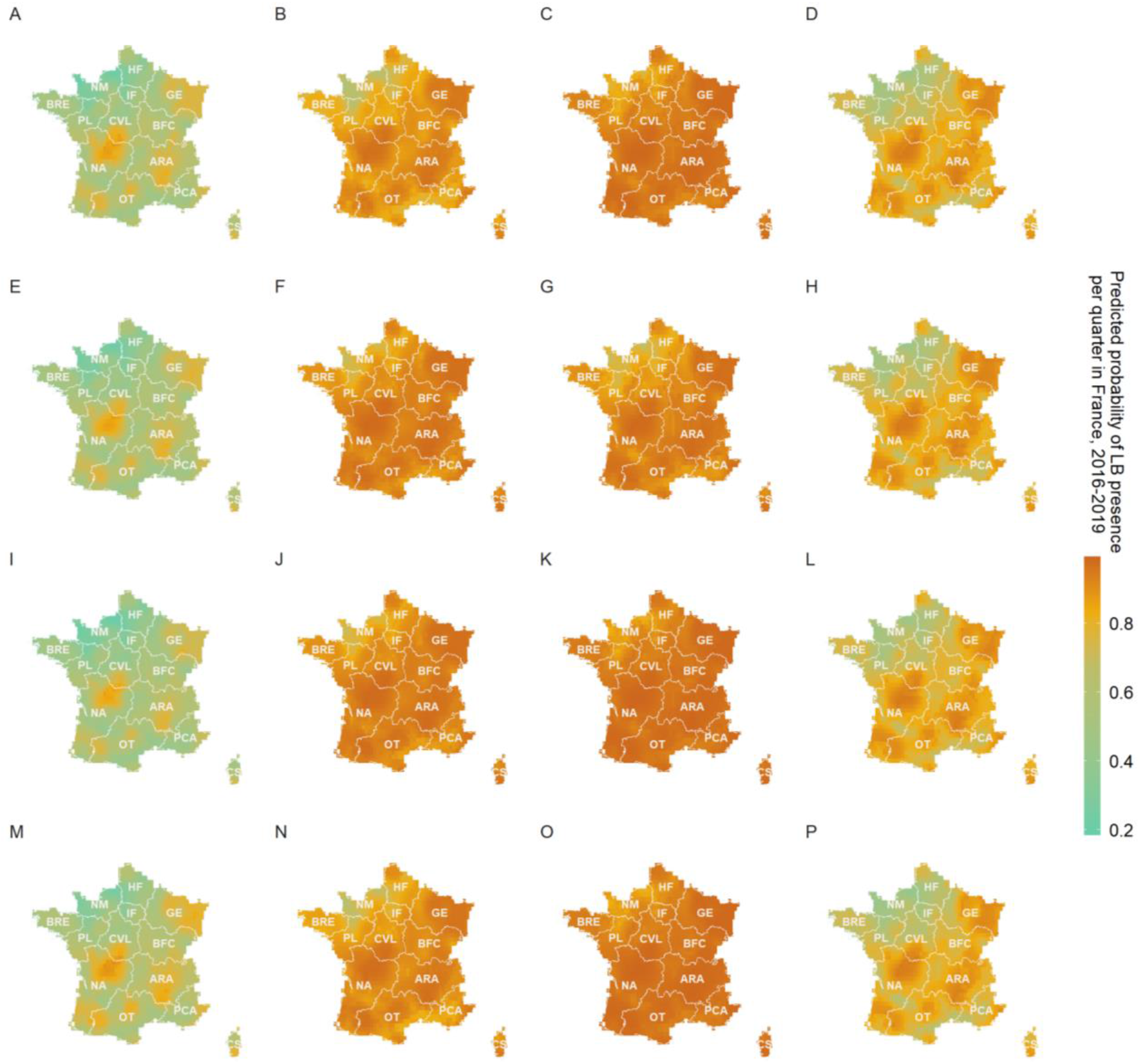
Predicted probability of LB presence in France per quarter, 2016-19. A-D for 2016, E-H for 2017, I-L for 2018, and M-P for 2019. Each column from left to right indicates the winter (January to March), spring (April to June), summer (July to September), and autumn (October to December) of each year. The darker orange color indicates a higher probability of LB presence, while the lighter green indicates a lower probability. A French shapefile with a resolution of approximately 0·2 × 0·2 decimal degrees (≈ 22km^2^) was used in this study (appendix pp5). For ease of interpretation, the name of each region is shown and referred to in the text as follows: HA=Hauts-de-France; NM=Normandie; IF=Île-de-France; GE=Grand Est; BRE=Bretagne; PL=Pays de la Loire; CVL=Centre-Val de Loire; BFC=Bourgogne-Franche-Comté; NA=Nouvelle Aquitaine; ARA=Auvergne-Rhône-Alpes; OT=Occitanie; PCA=Provence-Alpes-Côte d’Azur; CS=Corse.

Maps of the predicted seasonal incidence rate (/100,000 inhabitants) of LB for the period 2016-19 are presented in Figure 2A-2P. They exhibit geographical heterogeneity, seasonality and interannual variations. Over the four years, the seasons with higher LB incidence were spring and summer, with higher incidence reported in Grand Est (GE), Auvergne-Rhône-Alpes (ARA) and Nouvelle Aquitaine (NA) regions, echoing the results of the logistic model. The spatial patterns appeared similar in the spring of the years 2017 and 2018 (figures 2F and 2J), with slightly higher predicted incidences than in 2016 and 2019 (figures 2B and 2N). The summer patterns appeared similar across years, yet with higher incidence rates in 2018 and 2019 (figures 2K and 2O), compared to 2016 and 2017 (figures 2C and 2G), especially for the GE, ARA, and NA regions. In contrast, spatial patterns in autumn (figures 2D, 2H, 2L, 2P) and winter (figures 2A, 2E, 2I, 2M) remained similar over the four years, with slight intra-regional fluctuations.

**Figure 2.**
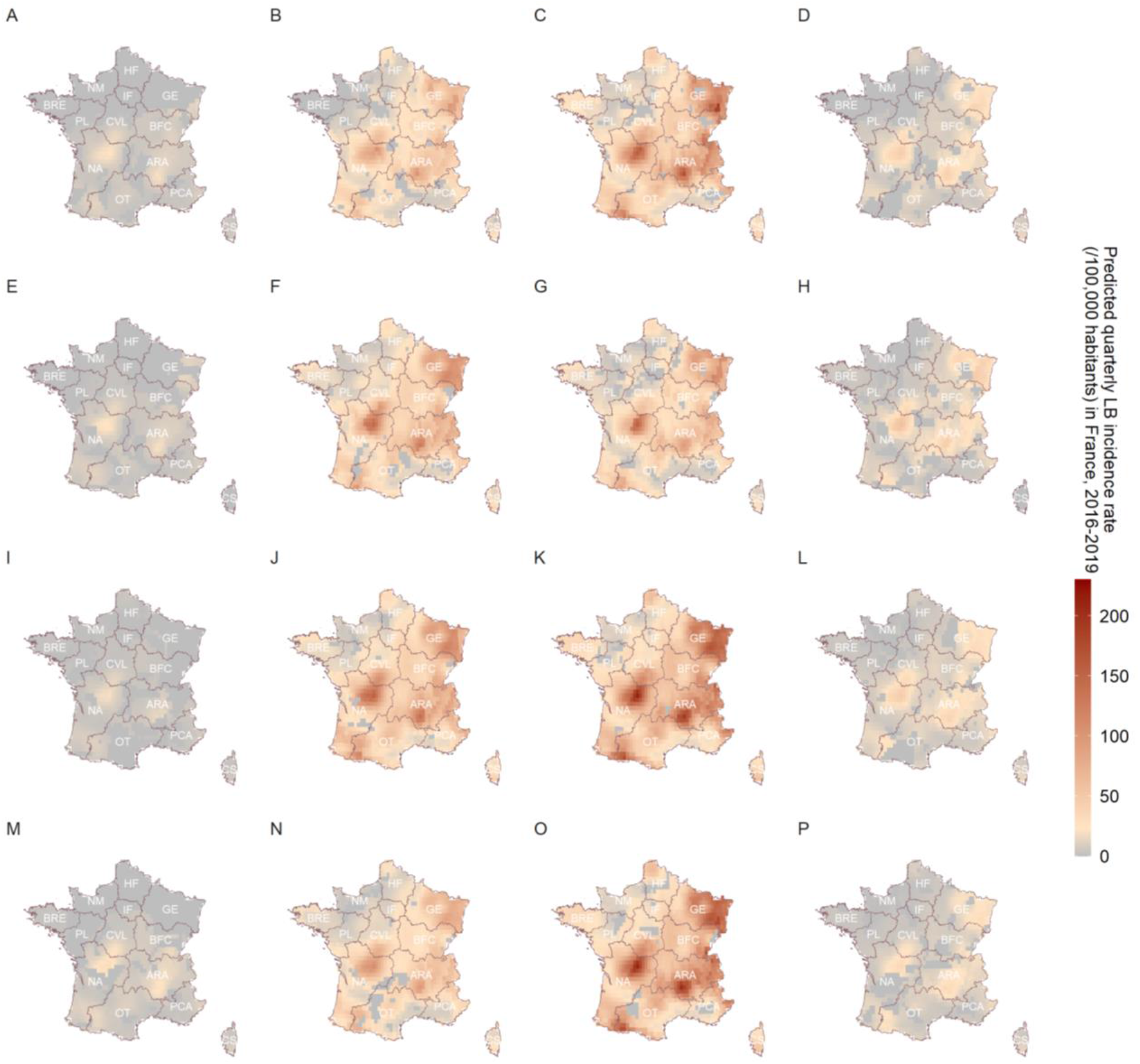
Predicted quarterly LB incidence rate (/100, 000 habitants) in France, 2016-19. A-D for 2016, E-H for 2017, I-L for 2018, and M-P for 2019. Each column indicates in turn the winter (January to March), spring (April to June), summer (July to September), and autumn (October to December) of each year. The darker red areas indicate a higher incidence of LB, while grey areas indicate no LB cases reported (ie, observations equal to zero that are excluded from the gamma model).

The forecast maps for 2020 are shown in Figure 3. We observed that the logistic (figures 3A-3D) and gamma (figures 3E-3H) model results have similar spatiotemporal patterns as in 2019. However, the incidence of LB increased in the summer of 2020 within sporadic grid cells in the eastern and western parts of ARA region (Figure 3G). We used the empirical distribution of the probability integral transform (PIT) to assess the predictive performance of the model and by comparing positive predicted values with observed data in 2020. The cross-validated PIT histograms showed a uniform distribution, indicating that the predicted distribution was consistent with the data (appendix pp 10). The other tested models are presented in appendix (pp 9). The variable number of rainless days was ultimately not included in the final model because its statistical relevance was not significant and also did not optimise the model selection results.

**Figure 3.**
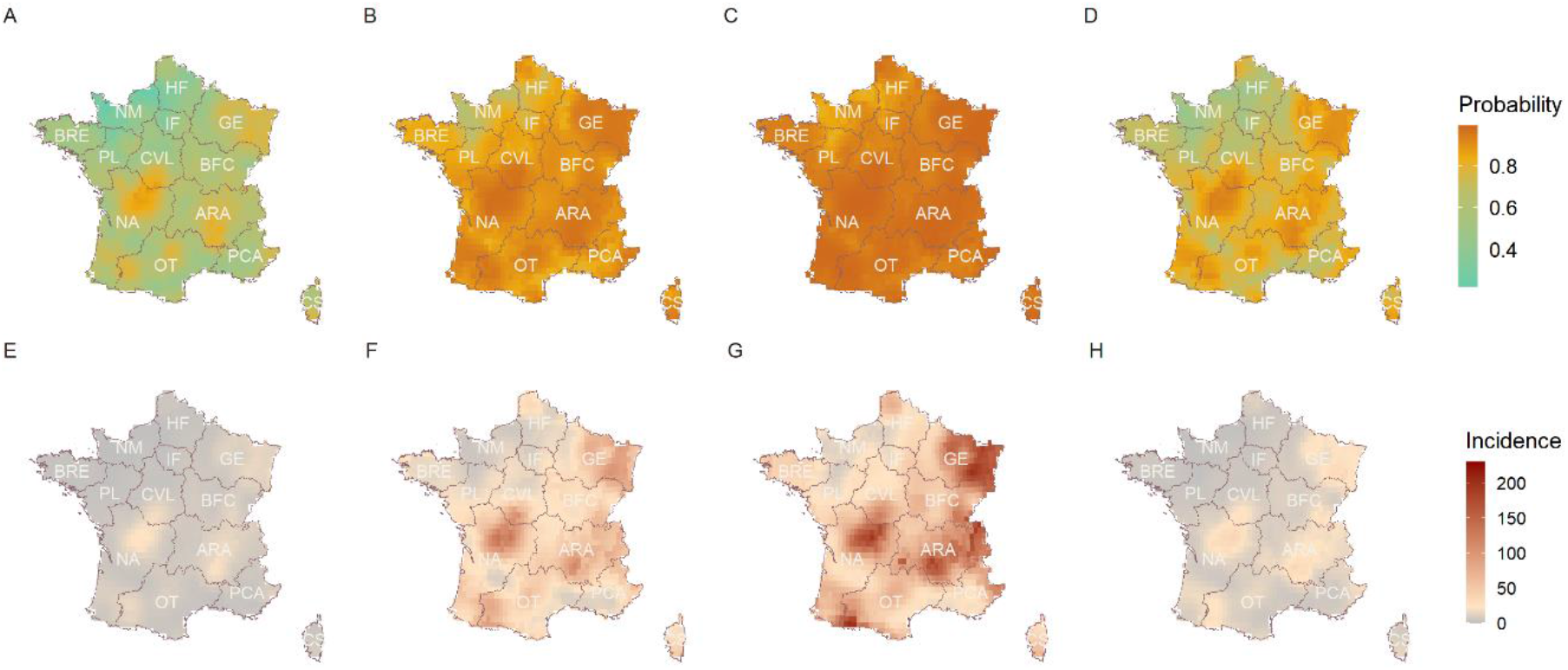
A-D Predicted probability of seasonal LB presence in France for 2020. E-H Predicted seasonal LB incidence rate (/100,000 habitants) in France for 2020.

## Discussion

We present the first national-level assessment of seasonal LB occurrence in Europe allowing to disentangle factors associated with LB presence and increased incidence. Our results provide evidence that seasonal LB presence was positively associated with higher vegetation index, which was used as a proxy for *Ixodes*, whilst increased incidence was associated with a high index of deer presence, mild seasonal temperature, moderate saturation deficit, and higher tick bite frequency. This model illustrates a spatio-temporal integrated analysis of meteorological, environmental, hosts, and anthropogenic factors for a zoonotic and vector-borne infection of major public health concern, and can be used as a reference model to be calibrated in other LB-affected areas.

One strength of our study lies in the use of kriging interpolation to obtain a continuous process of LB distribution in mainland France, allowing the inclusion of climatic and environmental variables at a fine resolution.^19^ In addition, we used a Bayesian two-part model and accounted for the effect of spatial correlation and seasonal variations, which attribute to reduce the structural random effects on parameter inference.^20^ Prediction and validation with 2020 data allowed to identify similar spatial and seasonal patterns across years, showing the robustness of our model. The Covid lockdown in France (March-May, 2020) was not expected to affect our validation dataset because humans acquire infection from tick bites, with major exposures occurring hereafter and LB cases peaking between May and August. Our prediction maps highlighted the seasonal pattern of LB occurrence, ie, spring and summer (April to September), and revealed the heterogeneous distribution of LB across and within regions by season and year in mainland France. Areas with a higher LB incidence burden were located in eastern (GE, BFC, and ARA), midwestern (NA) and southwestern (OT) France, in agreement with previous studies.^16^

NDVI value has been discussed in earlier studies as an important predictor of tick suitable habitat and questing nymph abundance.^23,30^ We incorporated NDVI into the model and found that areas and seasons with NDVI values greater than 0·6 were positively correlated with the occurrence of human LB, echoing the acarological studies. In addition, we introduced an indices of rodent species richness in the logistic model as a proxy for the extent of *Borrelia* infection. We found a negative correlation between LB presence and a greater number of rodent species, which is consistent with the results of a US study showing a weak negative correlation between the nymphal infection prevalence and species richness.^31^ There are several possible explanations for this. First, the available data provide only the number of species at each site; the exact species and density variation is unknown. More competent species but at low densities may not be able to maintain the local intensity of pathogen transmission.^32^ Second, the presence of more species also suggests that the local environment is friendly to small rodents and their predators and that coexisting other noncompetent reservoir hosts may have a diluting effect on tick infection rates.^32^ If larvae attach to these noncompetent hosts, nymphal infection rates are instead reduced. For further discussion, a nationwide field survey of species associated with *Borrelia* reservoirs and their densities should be necessary.

In addition, we found that increased LB incidence was associated with deer suitable habitat coverage of more than 80% of each grid area. The percentage of deer habitat laterally reflects the likelihood of deer presence and their abundance. Roe deer and red deer are known to be the primary productive hosts for adult female ticks and can maintain high tick populations in areas.^33^ As noncompetent *Borrelia* reservoir hosts, some studies have shown that ticks lose *Borrelia* infection after feeding on deer.^34^ However, others have suggested that an increase in overall tick density leads to an increase in the density of infected nymphs, which may negate the effects of diluting infection rates, resulting in increased LB risk.^35^ A long-term study in Norway also showed that high spatial and temporal densities of deer lead to an increased incidence of LB in humans, supporting our interpretation.^33^

Our findings also point to a positive correlation between areas with higher LB incidence and mild soil temperature (10–15°C) and moderate saturation deficit (3–5 mmHg). In our hypothesis, soil temperature in the previous season mainly influenced tick development, ie, attachment to other animal hosts for blood feeding to complete molting, which was indirectly related to human LB in the following season; whereas saturation deficit (a mixed indicator of temperature and humidity) in the current season mainly influenced tick’s human-seeking activity and was directly correlated with human incidence in that season. Optimal ground temperatures for tick activity have been shown to be between 13 and 15 °C, and most questing tick activity was found to occur between 2 and 7 mmHg, consistent with our model results.^26,27,36,37^ Thus, we speculate that larval tick hatch from eggs during mild springs and infest rodents acquiring *Borrelia* infection in endemic areas. Humans act as incidental hosts and are then exposed to these infectious tick bites during the nymphal and adult stages, resulting in an elevated risk of LB. In addition, the higher frequency of human outdoor activity during the summer months, which overlaps with tick habitat and activity periods, may also contribute to increased incidence.

The use of citizen-based health data is of rising interest in epidemiological research and has recently been applied to tick bite tracking in several countries.^13^ These citizen engagement-based data can be considered complementary to medical data, especially given the reporting bias and reporting delays in GP consultations, which may offer the potential for earlier detection of the onset of transmission. Here, CiTIQUE data allowed accounting for the spatial and temporal variations in the risk of human exposure to tick bites. Such data, assumed to capture at the same time human outdoor activity and tick encounter largely outperformed the number of rainless days (variable initially used as a surrogate to human outdoor activities) in improving model fit, highlighting the importance of such citizen-based research initiatives.

There are some limitations to consider. First, the national LB surveillance incidence used in this study was estimated based on cases reported by Sentinelles general practitioners (SGPs) in each department. Variations in the heterogeneous distribution, number, and frequency of reporting of SGPs may lead to some statistical differences. Second, the data collection from the national CiTIQUE programme is based on the general population. Uneven public awareness of ticks and differences in the frequency of application use across regions may affect tick bite reporting. In addition, CiTIQUE data is available after July 2017 and we calculated seasonal averages by department for 2016–19, which may also contribute to some uncertainty in the results. Third, conceptually, the zero values in the two-part model are considered to be true zeros. We observed that most zero values occurred in winter and autumn, consistent with the epidemiological characteristics of LB occurrence, but zero values were also generated in spring and summer in some high-risk areas. Therefore, zero values in our data could be a result of the absence of LB cases or lack of SGP reporting. This also emphasizes the need for continuous and enhanced surveillance and improved quality of data to reduce uncertainty in parameter inference. Furthermore, in our analysis, the effects of meteorological factors were assumed to have an impact on tick development and activity. Yet, we acknowledge that they also affect the reproduction rate and activity of other potential hosts (especially small mammals), as well as their food resources (eg, tree seeds). Meteorological factors may also be associated with human propensity to be outdoors, particularly when weather favours outdoor recreational activities and overlaps with weekends, holidays, and summer vacations, potentially generating more reports of human tick bites. Given that some risk factors have now been identified, further exploration of the complexity and dynamics of human LB incidence should be complemented by using mathematical models.

## Data Availability

All data produced in the present work are contained in the manuscript

https://cds.climate.copernicus.eu.

https://land.copernicus.eu.

https://openhealthdata.

## Contributors

WF, CB, and RM designed the study. WF, RD, and RM completed the modelling and statistical analysis. WF, CB, JF, AS, JD, PFK, RD, BJ, and RM interpreted and discussed the results. WF drafted the paper. CB, JF, AS, JD, PFK, RD, BJ, and RM contributed to the review of the paper. WF and RM completed the paper. All authors contributed to data interpretation, and reviewed and approved the final version manuscript. All authors had full access to all the data in the study and had final responsibility for the decision to submit for publication.

## Acknowledgments

WF is funded by a Sorbonne University PhD fellowship, JD is supported by a grant overseen by the French National Research Agency (ANR) as part of the «Investissements d’Avenir» program (ANR-11-LABX-0002-01, Lab of Excellence ARBRE). We are grateful to the Réseau Sentinelles, for providing the data, and all the Sentinelles general practitioners participated in the monitoring activities in France. We are also grateful to the CiTIQUE team members who initiated and are running this participatory research: Jean-François Cosson and Gwenaël Vourc’h from INRAE; Annick Brun-Jacob from Lorraine University; Irene Carravieri, Julien Marchand and Cyril Galley from CPIE Nancy-Champenoux. Finally, we thank all citizens who participated to CiTIQUE.

## Data sharing

The meteorological data used can be downloaded from https://cds.climate.copernicus.eu. Vegetation data can be downloaded from https://land.copernicus.eu. Animal host data can be download from https://openhealthdata. Requests for tick bite information can be addressed to. Requests for LB surveillance data can be addressed to. The model code can be downloaded from https://github.com/wenfu27/TPBM_LB.

